# A strategy to assess spillover risk of bat SARS-related coronaviruses in Southeast Asia

**DOI:** 10.1101/2021.09.09.21263359

**Authors:** Cecilia A. Sánchez, Hongying Li, Kendra L. Phelps, Carlos Zambrana-Torrelio, Lin-Fa Wang, Kevin J. Olival, Peter Daszak

**Author notes:** Corresponding author EcoHealth Alliance, 520 Eighth Ave., Suite 1200, New York, NY 10018.

## Abstract

Emerging diseases caused by coronaviruses of likely bat origin (e.g. SARS, MERS, SADS and COVID-19) have disrupted global health and economies for two decades.

Evidence suggests that some bat SARS-related coronaviruses (SARSr-CoVs) could infect people directly, and that their spillover is more frequent than previously recognized. Each zoonotic spillover of a novel virus represents an opportunity for evolutionary adaptation and further spread; therefore, quantifying the extent of this “hidden” spillover may help target prevention programs. We derive biologically realistic range distributions for known bat SARSr-CoV hosts and quantify their overlap with human populations. We then use probabilistic risk assessment and data on human-bat contact, human SARSr-CoV seroprevalence, and antibody duration to estimate that ∼400,000 people (median: ∼50,000) are infected with SARSr-CoVs annually in South and Southeast Asia. These data on the geography and scale of spillover can be used to target surveillance and prevention programs for potential future bat-CoV emergence.

## Introduction

Emerging coronaviruses (CoVs) of wildlife origin have significantly disrupted global health security and economies during the last two decades^1,2^. Severe acute respiratory syndrome (SARS) and Middle East respiratory syndrome (MERS) CoVs caused significant human morbidity and mortality in 2002 and 2012 respectively^3,4^. Swine Acute Diarrheal Syndrome CoV caused substantial mortality in pigs in southern China during 2016 and 2019^5,6^. The emergence of SARS-CoV-2 in 2019 led to the current COVID-19 pandemic that has caused millions of cases and deaths, with economic loss likely to be in the tens of trillions of US dollars^2,7^. Efforts to increase preparedness and improve surveillance for emerging coronaviruses therefore represent a priority for global health programs^8^.

Phylogenetic analysis suggests that SARS-CoV, MERS-CoV, SADS-CoV, and SARS-CoV-2 originate within CoV lineages from bat reservoir hosts^6,9-11^. The initial spillovers of SARS and MERS into human populations are thought to have occurred via intermediate hosts (palm civets and dromedary camels, respectively^12,13^). However, the role of civets in the emergence of SARS is uncertain, and other bat SARSr-CoVs can directly infect human cells, including airway epithelial cells, and thus have potential to spill over directly from bats to humans^14-16^. It is uncertain what proportion of bat SARSr-CoVs can infect people either directly or indirectly (via an intermediate or amplifier host). However, serological evidence of prior infection with SARSr-CoVs in communities living near bat populations in China prior to the emergence of COVID-19, including in people who reported no contact with SARSr-CoV intermediate hosts, suggests direct bat-to-human transmission might occur in some regions^17,18^. Direct bat-to-human spillover events may occur more frequently than has been reported, but go unrecognized because they cause mild symptoms, cause symptoms that are similar to other infections, result in small numbers of cases, or lack sustained chains of human-to-human transmission. However, every wildlife-to-human spillover event represents an opportunity for viral adaptation permitting human-to-human spread^19-22^. Quantifying the extent of these undetected spillovers could therefore be important to identifying risk of future epidemics or pandemics.

Surveys of bats in China have revealed high diversity of SARSr-CoVs, and often high prevalence (5-10%) in rhinolophid and hipposiderid species that are widely distributed, abundant, resilient to habitat perturbation and are synanthropic (have contact and often interactions with human populations)^23-25^. Many of the bat species and genera known to harbor these β-CoVs occur in Southeast Asia, a hotspot of bat diversity with 441 species reported, 115 (around a quarter) of which are rhinolophids or hipposiderids^26^. While some related β-CoVs have been reported outside China (including within Africa), phylogenetic analyses of SARSr-CoVs indicate that regions of south China and parts of neighboring countries (Myanmar, Lao PDR, and Vietnam) act as areas of evolutionary diversification with high diversity of these viruses. However, sampling in China has been far more intense than in nearby Southeast Asian or South Asian countries, and diversity of these viruses is also likely high in these regions^23^. Furthermore, many of these less well-sampled countries are undergoing dynamic social and environmental changes correlated with zoonotic emergence (e.g. rapid human population growth, movement of rural residents to urban centers, extensive wildlife farming and trade, and rapid land conversion from pristine forested habitats to agricultural monocultures), and thus might represent hitherto unreported hotspots for coronavirus spillover risk^23,27-32^.

In this study, we use host distribution modeling, ecological, and epidemiological data to estimate the geographic distribution of SARSr-CoV exposure risk, and likely rate of unreported zoonotic spillover in China, South and Southeast Asia. Our results provide the most detailed estimates to date of the distribution and species richness of bat hosts of SARSr-CoVs, and suggest that human exposure to and spillover of SARSr-CoVs may be substantially underestimated. Our approach provides proof of concept for systematic risk assessment of zoonotic spillover, and a strategy to identify key geographic areas that can be prioritized for targeted surveillance of wildlife, livestock, and humans. Given the challenges of identifying the origins of COVID-19 and pathways by which SARS-CoV-2 spilled over to people^33,34^, this approach may also aid efforts to identify the geographic sites where spillover first occurred.

## Results

We assembled a list of 23 known SARSr-CoV bat host species, mainly members of the *Rhinolophidae* and *Hipposideridae*, occurring in the geographic region of interest (Table S1). We derived an area of habitat (AOH) for each species (Fig. 1, also see rationale for using this approach in Methods), by refining the IUCN geographic range of each species using habitat suitability, elevation limits, and the boundaries of a broadly defined region including parts of South Asia, China and Southeast Asian countries. Removing unsuitable areas of bat occurrence within the IUCN geographic ranges highly improved assessment of species distribution. For example, the reduction in area from the original IUCN geographic range to the more refined AOH for each species ranged from 42% (*Rhinolophus malayanus*) to almost 100% (*R. hipposideros*), with a median of 68% reduction (Fig. S1). Notably, the reduction in area was 55% for *R. rex*, the only species in our list assessed as endangered by the IUCN Red List.

**Figure 1.**
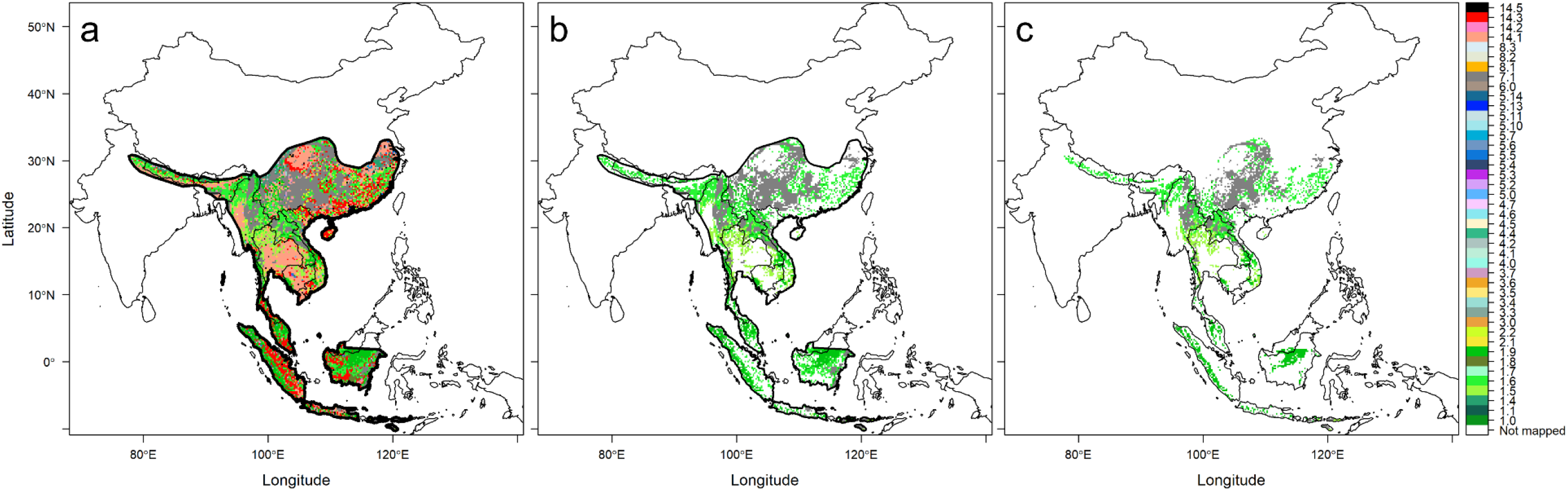
Representative illustration of IUCN range refinement by habitat type and elevation limits to create area of habitat (AOH). **a** The IUCN range of *Rhinolophus affinis*, outlined in black, is overlaid on a map of terrestrial habitat types, and the habitat map is restricted to only the species range. **b** Only suitable habitat types within the species range are retained. **c** After further restricting habitat by the elevation limits of the species, the remaining habitat represents the AOH of *R. affinis*. The color scheme follows that of ^72,73^, with the addition of gray used for habitat type 7.1 (caves; carbonate rock outcrops used as proxy). Names of habitat types are found in Fig. 2b and Table S1.

We validated species AOHs using cleaned occurrence records downloaded from the Global Biodiversity Information Facility (GBIF; see Methods for details of data cleaning and validation). After data cleaning, no occurrence records remained for three species (*Hipposideros pratti, R. ferrumequinum*, and *R. hipposideros*), while 1-179 (median = 13) occurrence points remained for all other species (Table S2). In validating each species’ AOH with GBIF occurrence points, we found among species with ≥1 occurrence point, the median percent of points (buffered by 5 km) that overlapped a species’ AOH was 64% (Table S2). Among species with at least 30 occurrence points (*n* = 7), the median overlap was 84%. Overlap was >80% for five species: *R. creaghi* (31/31, 100%), *R. malayanus* (8/8, 100%), *Chaerephon plicatus* (45/53, 85%), *H. galeritus* (37/44, 84%), and *H. larvatus* (41/49, 84%).

The size of individual species AOHs varied widely, but generally, the largest AOHs encompassed the most people (Fig. 2a). For example, the AOH of *R. affinis* was the largest of all species, covering ∼1.9 million km^2^, and it encompassed ∼132 million people (2nd highest of any species). Two species had AOHs with more limited area but relatively high human population density: *Nyctalus leisleri* (∼262 people/km^2^) and *R. stheno* (∼226 people/km^2^; Fig. 2a, Fig. S2). Within species AOHs, forest habitats and ‘carbonate’ (limestone) rock outcrops (used as a proxy for cave distribution) typically comprised the largest proportion of suitable habitat (Fig. 2b). Two species (*R. stheno* and *R. malayanus*) had high proportions (>50%) of artificial habitats, including plantations and arable land.

**Figure 2.**
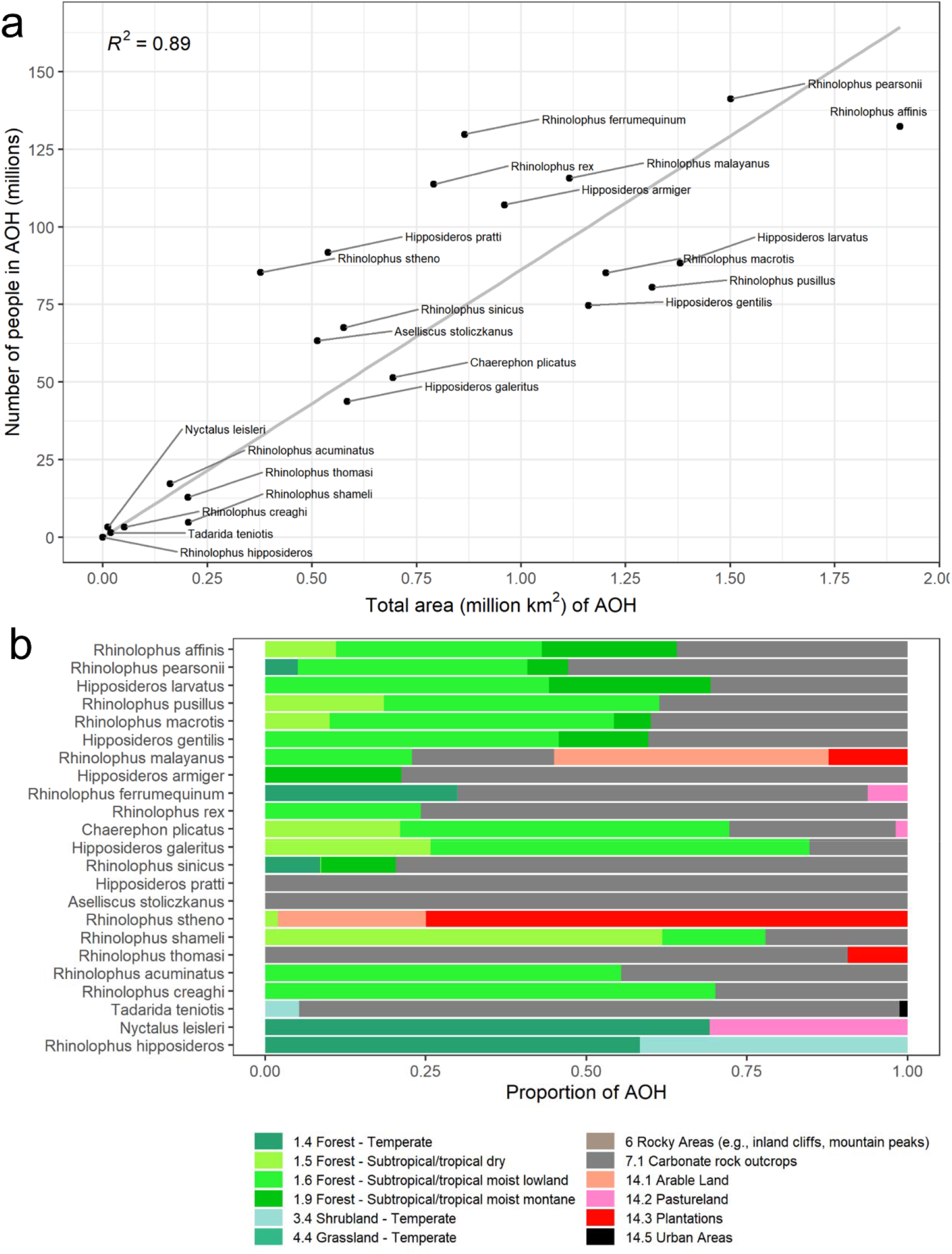
Relationships among AOH size, number of people, and habitat proportions. **a** Scatterplot showing the total number of people living in each AOH versus the total area, for each SARSr-CoV bat host species. A best-fit line was fit through the origin, for which the R^2^ is displayed. **b** Proportion of each habitat type within species AOHs. Species are listed in order of decreasing AOH size. Color scheme follows that of Jung and colleagues^72,73^, with the addition of gray for caves (carbonate rock outcrops used as proxy).

The consensus area of all SARSr-CoV bat host species, created by mosaicking the 23 species AOHs, comprised ∼4.5 million km^2^. We calculated that ∼478 million people live within this consensus area, which covered most of Lao PDR, Cambodia, Thailand, Vietnam, Nepal, Bhutan, peninsular Malaysia, Myanmar, southeast China, and the western islands of Indonesia (Fig. 3a). Bat species distribution was patchier in India, Sri Lanka, East Malaysia, and the Philippines. Species richness ranged from 1-14 species, with the highest richness of SARSr-CoV bat host species in southern China, eastern Myanmar, and northern Lao PDR (Fig. 3a). When we visualized areas with both high host richness and large human populations (‘relative spillover risk’), southern China remained a hotspot, while other areas emerged as important because of their high human population sizes (e.g. Java, parts of northern India; Fig. 3b).

**Figure 3.**
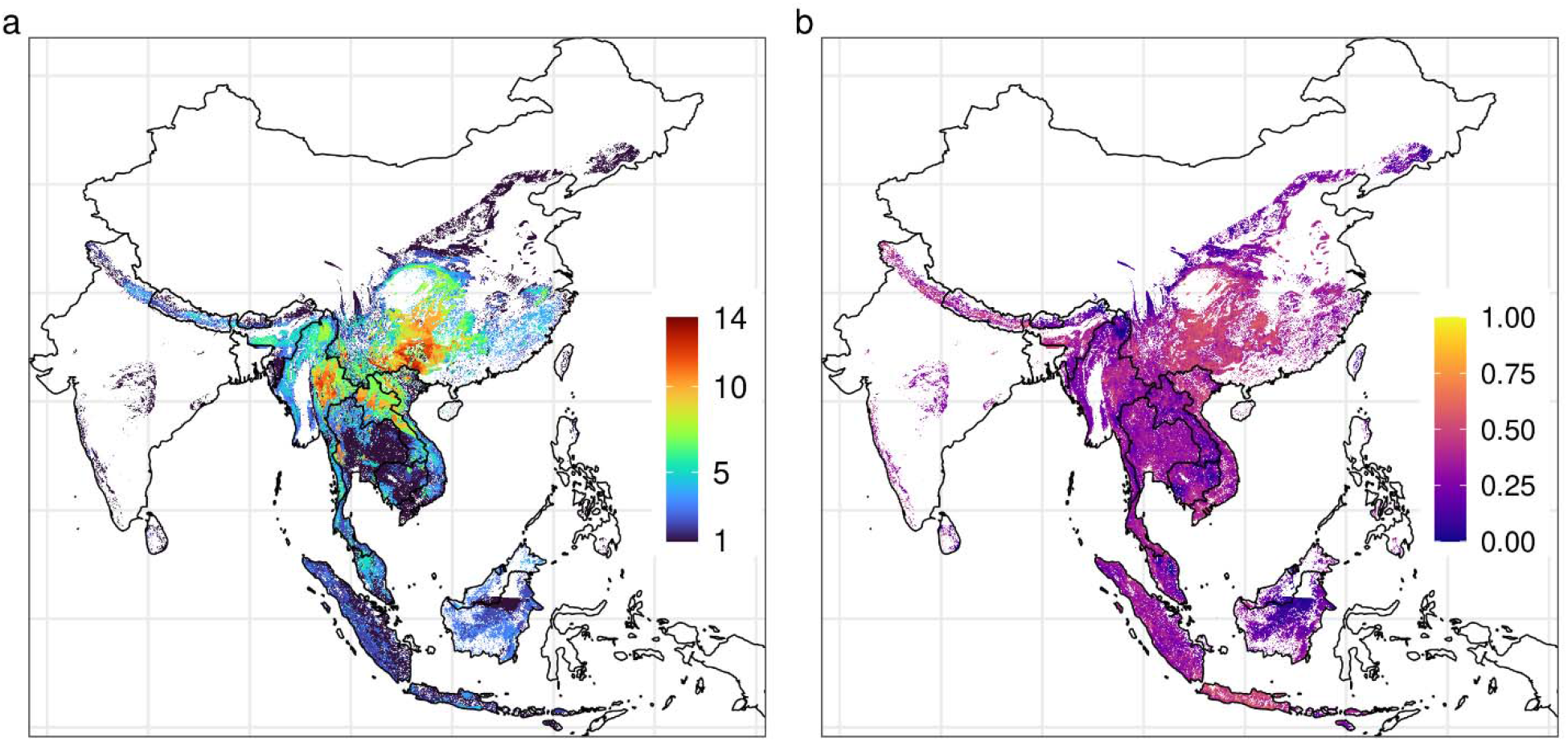
Hotspot maps of Southeast Asia. **a** Species richness of SARSr-CoV bat host species in Southeast Asia. This map was created by overlaying AOH maps for all 23 SARSr-CoV bat host species. **b** Bat host species richness * human population count, or “relative spillover risk”. Values were log(x + 1) transformed and then normalized to a 0-1 scale.

After calculating the number of people living in the consensus area of SARSr-CoV bat host species, we further incorporated data from the literature on bat-human contacts, human SARSr-CoV seroprevalence, and SARS antibody duration to estimate the extent of hidden bat-human spillover of SARSr-CoVs in Southeast Asia (see Supplemental Material for details of probabilistic risk assessment methods). We estimated that within the consensus area of SARSr-CoV bat host species, an average of 407,422 people (median: 53,225; range: 1 - 35,632,228) are infected with SARSr-CoVs each year in Southeast Asia (Fig. S3). Sensitivity analyses indicated that the probability of antibody detection given contact with a bat, and the probability of contact with a bat, contributed most to variance in the outcome (Fig. S4). In the process of identifying datasets of value for each step of the probability analysis, we also assessed datasets that are not available or for which quality could be improved, and would likely provide more accurate assessments of risk (Fig. 4).

**Figure 4.**
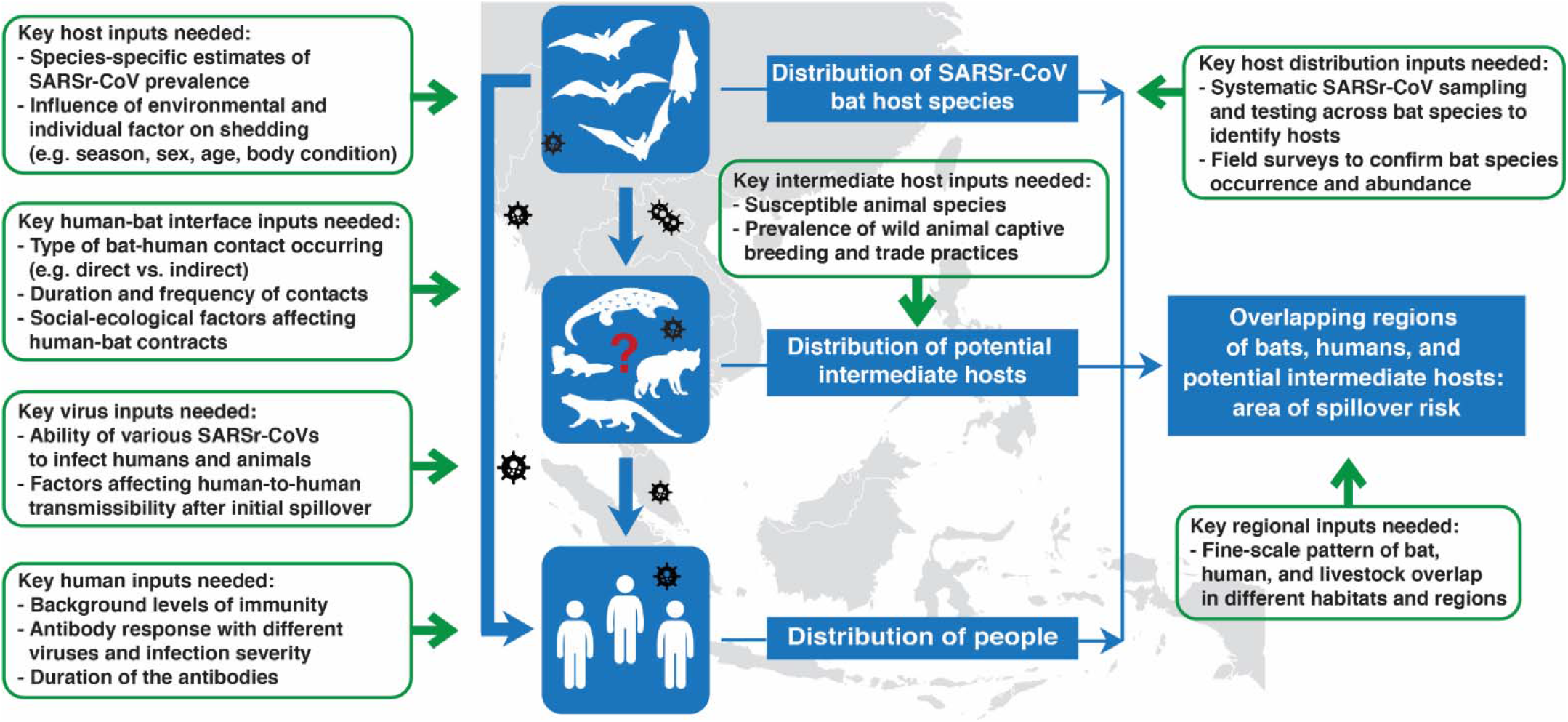
Conceptual figure depicting key data inputs to better estimate spillover of SARSr-CoVs from bats to humans, and gaps in the available data.

## Discussion

Our paper reports an analytical framework to assess SARSr-CoV spillover risk in a region that includes the site of the first spillover of SARS-CoV, and likely of SARS-CoV-2. By incorporating data on biologically plausible host distributions for all bat species known to harbor SARSr-CoVs in South and Southeast Asia, on human population counts, on behavior of people in the region that drives bat-human contact, and on seroprevalence in people and duration of antibody persistence, we are able to provide detailed maps of potential bat-to-human SARSr-CoV spillover hotspots, as well as estimates of the number of people infected by this viral group annually. Our approach may assist in better identifying regions for targeted surveillance of local communities at risk of spillover, for conducting viral discovery programs to identify novel bat-CoVs, for COVID-19 origins tracing, and for estimating human surveillance targets to identify spillover events earlier and more accurately. All of these are key goals for pandemic preparedness and prevention^35-38^, and if used to target future surveillance and disease control, may help to reduce the risk of future COVID-like outbreaks.

Our analysis identifies regions in southern China, northeastern Myanmar, Lao PDR, and northern Vietnam as having the highest diversity of SARSr-CoV bat host species. These hotspots of SARSr-CoV bat reservoir host diversity may be particularly fruitful sites for viral discovery of novel SARSr-CoVs, assuming that viral diversity scales with host species diversity^39^. This finding supports conclusions from prior phylogenetic analyses that particularly diverse SARSr-CoV lineages are found in southern China, and that it may be a center of evolutionary origin for this group of β-CoVs^23^. It also helps explain the recent identification of multiple strains of SARSr-CoVs in southern China^32,40,41^ and southeast Asian countries^31^, despite small sample sizes. It suggests that the less intense sampling in countries bordering southern China has led to an underestimate of the diversity of these viruses there. Given that the bat species known to host the closest relatives of SARS-CoV-2 are found in this region, these maps may also guide efforts to identify the viral clade from which a progenitor virus emerged^23,32,34^. The map of bat SARSr-CoV host diversity combined with human population density points to hotspots of bat-to-human exposure risk (and potential for human-to-human spread) in southern China and bordering countries, but also in the populous regions of Indonesia. This map may be useful in targeting surveillance to identify SARSr-CoV spillover events in people, including syndromic surveillance for SARS- or COVID-like respiratory disease in communities within these hotspots, as proposed previously^18,24,35,39^. It may also provide guidance for studies of where initial spillover of the progenitor of SARS-CoV-2 may have occurred, although epidemiological and other data suggest this would most likely have been in China or neighboring countries^33,34^.

Our estimate that an average of ∼400,000 (median ∼50,000) people are infected with SARSr-CoVs each year in Southeast Asia suggests that bat-to-human SARSr-CoV spillover is common in the region, and is undetected by surveillance programs and clinical studies in the majority of cases. While the data suggest significant levels of exposure, many of the diverse viral strains that infect people in the region each year may not be able to replicate well in people, cause illness, or be transmitted sufficiently among people to cause an outbreak. This has been shown in theoretical models of disease emergence^21,42^, and supports earlier evidence from studies of non-human primate virus spillover^43^, that cross-species transmission of novel animal-origin viruses is not the rate-limiting step in pandemic viral emergence. However, given the relatively large number of people likely to be infected each year with bat-CoVs, it is plausible that illnesses or clusters of cases due to novel bat-CoV infection occur regularly within the region, and are either not reported, or otherwise missed by clinical surveillance. Evidence of underreporting has been demonstrated for other bat-origin viral infections. For example, targeted syndromic surveillance of encephalitis patients in a small number of clinics in Bangladesh showed that Nipah virus causes outbreaks annually with an overall mortality rate of ∼70%, despite it only recently being reported from the country^44^. Efforts to increase surveillance for novel SARSr-CoVs (and other bat-origin viruses) in clinical cohorts, particularly using syndromic surveillance, may identify the rate of missed cases, and pre-empt larger scale outbreaks. Estimating the true rate of spillover of previously unknown, potentially zoonotic animal-origin viruses is difficult without serological or genomic surveillance data. For most viruses, the duration of infection in humans is relatively short, e.g. the infectious period for COVID-19 is 2-3 weeks^45^, and if spillover is rare, PCR surveillance is unlikely to give valuable data on spillover rates due to lack of positives. One exception is for viruses with long infectious periods such as lentiviruses and some retroviruses, and a previous genomic surveillance study of wildlife hunters in Africa was able to find 10/1099 (0.9%) prevalence of non-human primate origin simian foamy viruses^46^. Because detectable antibodies or T cell responses are more long-lived, serological surveys or activated T cell testing represent more valid strategies to estimate spillover rates.

Our calculation of undetected spillover represents the first published attempt to identify the rate of spillover of novel coronaviruses from bats to people. It relies on a number of input variables and we attempted to account for uncertainty by assigning a probability distribution to each variable, but recognize that further data would likely help refine estimates of SARSr-CoV spillover rates. Perhaps most critical is a lack of information on the role of intermediate hosts in the emergence of SARSr-CoVs. It has been postulated that civets and other commonly farmed and traded mammalian species played a role in the emergence of SARS-CoV by acting as efficient amplifier hosts within wildlife farms and markets^12,47-50^. SARS-CoV-2 can infect civets, raccoon dogs, and other mammals commonly farmed and traded for food in the region^51,52^. SARS-CoV-2 has also caused significant outbreaks in animals bred for fur (e.g. mink, raccoon dogs) that have in some cases led to transmission to people^53,54^. These and other data led an international team to conclude that the most likely pathway for COVID emergence was from bats to people through a farmed mammalian intermediate host^33^. Accurate data are not available on the number of wildlife farms and potential intermediate hosts bred each year, or on the market systems that supply live animals into cities within China and Southeast Asia. It was estimated that 14 million people were employed in the wildlife farming system within China alone during 2016, in an industry worth $77 billion annually^55,56^. Additionally, some of these potential intermediate hosts occur naturally in the region (e.g. pangolins, civets), and other livestock species that are extremely common (e.g. pigs, cattle, rabbits) are susceptible to SARSr-CoVs either naturally or experimentally^52^. Spillover of SARSr-CoVs may therefore be substantially skewed to people who have high exposure to these species, and this would likely have been missed in the relatively small serological surveys upon which our analyses are currently based. Thus, better estimates of the role of farmed and traded intermediate hosts are likely to substantially increase the estimated spillover rates of SARSr-CoVs across the region.

Other data could improve our estimates. These include expanded survey data on the background seroprevalence of bat SARSr-CoVs in people across different populations and geographies, analysis of how specific risk behaviors correlate with seroprevalence, and studies of how serological titers and duration of antibody persistence relate to severity of infection. We estimated bat-human contact using data from previous human-animal contact surveys and ethnographic investigations, but contacts likely vary widely across the region due to different cultural practices and traditions^57,58^. The type of contact may be critical to assessing the risk of transmission (e.g. hunting and butchering vs. living near a bat colony), and some types of contact may be unrecognized and therefore unreported (e.g. exposure to feces or urine on surfaces). Some of the contact data we identified in our systematic literature search (see Supplemental Material for details) were from populations targeted because of known high risk of bat contact due to occupational or environmental exposure and are likely upwardly biased due to non-random study design. Expanded bat viral survey data would also likely improve our estimates, first by obtaining more contemporary, accurate species presence records, and also by identifying previously unknown SARSr-CoV hosts, improving species-specific estimates of SARSr-CoV prevalence, and clarifying how environmental (e.g. location, season, year) and host trait (e.g. sex, age, body condition) factors affect viral shedding^59-62^. Finally, it is also possible that taxonomic errors have occurred in the data we analyzed. Taxonomic standards in viral discovery and surveillance studies vary widely, and point to a need for better taxonomic training of field teams, standardized DNA ‘barcoding’ of hosts, collection of voucher specimens, and closer collaboration among disease ecologists, virologists, field biologists and taxonomists^63,64^.

Our refinement of species ranges has likely produced the most accurate contemporary picture of SARSr-CoV bat host species occurrence to date, and demonstrates substantial range modification for many species, reflecting rapid land use change throughout the region. The AOHs we produced may be useful for targeting surveillance to key species. For example, only a few species AOHs contained a sizable proportion (≥ 25%) of one or more artificial habitats (Fig. 2b): *R. malayanus* (arable land, plantations), *R. stheno* (plantations, arable land), and *N. leisleri* (pastureland). These species may be more likely to come into contact with humans and could represent a particularly important spillover risk considering that global analyses demonstrate zoonotic host diversity increases in areas with anthropogenically modified habitats^65^. Cave habitats were classified by the IUCN as suitable for nearly all species in our analyses, and carbonate rock outcrops (used here as a proxy for caves) made up a large proportion of species AOHs. Visiting caves, collecting guano, and using bat guano in crop production are likely particularly high-risk activities given these findings^66-69^. Our validation process indicated good agreement between species AOHs and GBIF occurrence data, suggesting that the maps we generated accurately reflect species presence for occurrence records collected after 1990. Limitations of AOH maps^70^ include the potential inaccuracy of the IUCN species ranges^71^, habitat suitability assignments, and elevation limits. Additionally, the map of terrestrial habitat types we used in our analyses^72,73^ did not include caves, an important habitat type for many bat species. We used carbonate rock outcrop data as a proxy for cave distribution and this could be ground-truthed.

Our analytical framework provides a strategy that has potential for improving preparedness for emerging diseases and pandemic risk. It has produced maps that can be used to conduct more cost-effective field surveys for viral discovery programs, and estimates of spillover rates that can guide targeted human surveillance to identify clusters of cases of a new CoV infection earlier and help prevent spread. It may also give vital guidance for efforts to identify the reservoir hosts of the SARS-CoV-2 progenitor, and the sites of COVID origins or first emergence^34^. Our analysis pipeline and framework are based on open-source code and can therefore serve as a resource to update and modify spillover risk maps and estimates as new data become available. These could include serological surveys in people using new diagnostic assays that can detect virus-specific neutralizing antibodies to differentiate COVID-19 variants, vaccine strains, and different clades of bat SARSr-CoVs^74^, or data from ethnographic surveys that identify changes in bat-to-human contact as habitats are increasingly modified or laws to reduce hunting or wildlife trade reduce consumption are enforced. Finally, our framework can be rapidly adapted for spillover risk assessment of other viral groups, such as the HKU-2/SADS-CoV α-CoVs that have recently been found able to infect primary human airway epithelial cells *in vitro*, and therefore pose a heightened spillover risk, or any of the other ∼25 viral families that include known zoonoses^39^.

## Methods

### 1. Compilation of SARSr-CoV bat host data

We identified all bat species from which molecular evidence of SARSr-CoV infection had been reported, and for which associated sequence confirmation data were available. We supplemented a recently compiled list^75^ with hosts listed in other more recent publications^23,30,31,41,76-78^. We removed duplicate records and hosts only identified to genus level. We considered *Rhinolophus paradoxolophus* to be a subspecies of *R. rex*^79^. We excluded *R. monoceros* because, although recent work has retained it as a distinct species^80^, no recent assessment of the species has been published by the IUCN Red List of Threatened Species. *Hipposideros pomona* and *H. gentilis* were split in 2018, with *H. pomona* restricted to a small area in southern India while *H. gentilis* is broadly distributed across Southeast Asia^81^. Although *H. pomona* was listed as a host species in ^23^ from field sampling in China, we therefore substituted *H. gentilis* in our analyses. We chose to not include several bat species that were predicted to be SARSr-CoV reservoirs using host-trait or species network models^82,83^, as these predictions remain unvalidated, and vary in reliability across models.

We restricted the compiled list of SARSr-CoV bat hosts to those with geographic distributions either partially or entirely within Southeast Asia, and included the following countries and administrative regions: Bangladesh, Bhutan, Brunei, Cambodia, China, Hong Kong SAR, Macao SAR, India, Indonesia, Lao People’s Democratic Republic, Malaysia, Myanmar, Nepal, the Philippines, Singapore, Sri Lanka, Thailand, Timor-Leste, and Vietnam. See Table S1 for the finalized list of SARSr-CoV bat hosts. Note that while we use the term ‘Southeast Asia’, the region we considered has broader range than is used for political definitions of the region and includes China and parts of South Asia, due to the extensive range of some of the bat host species.

### 2. Calculation and validation of host area of habitat

All analyses were performed in the R statistical environment v. 4.0.3 (R Core Team 2020). To derive a biologically realistic area of habitat (AOH) within our Southeast Asia region for each SARSr-CoV bat host reservoir species, we refined species ranges by incorporating data on habitat suitability and elevation limits^70^. We used an AOH approach rather than ecological niche modeling (ENM) because ENMs rely on presence data, and this was difficult to find within GBIF, particularly when we excluded older records that may not reflect current distributions. Secondly, ENMs have the potential to overestimate range sizes and are thus not useful for identifying more precisely areas for surveillance, for example. To derive the AOH for each species, we downloaded its geographic range from the IUCN Red List of Threatened Species^84^ (last update: March 25, 2021), overlaid it onto a raster map of terrestrial habitat types^72,73^, and selected areas of the map that occurred within its range (Fig. 1a). We then selected areas of suitable habitat (as determined by IUCN) for each species (Fig. 1b, Table S1; see Supplemental Material for further details). Finally, we overlaid elevation data (Shuttle Radar Topography Mission data obtained with the getData function of the “raster” package^85^) onto each species’ range and habitat map and selected areas that fell within a species’ elevation limits (Fig. 1c; Table S1); these remaining areas represented the species’ AOH. Using the “raster” package, we calculated the size (in km^2^) of each species’ AOH and compared it to the size of its original IUCN range. Note that all analyses were restricted by the regional boundaries of Southeast Asia as defined above.

We validated the AOH of each species using occurrence data downloaded from the Global Biodiversity Information Facility (GBIF) using the “rgbif” package^86,87^. We cleaned GBIF data by removing records with inaccurate or imprecise coordinate data (i.e. no coordinates, identical longitude and latitude, coordinates of 0, coordinate uncertainty > 35 km, coordinates in the ocean, country–coordinate mismatch), records from areas outside our geographically defined region, records of absence rather than presence, and records with unreliable data sources (e.g. fossil specimen, literature, unknown). We removed records within 5km of country capitals, within 1km of country/province centroids, and within 100m of biodiversity institutions. We removed records before 1991, as older records tend to have less precise location data^88^ and could reflect species ranges that have since shifted. Within each species, we removed records with duplicate coordinates. Cleaning was facilitated with the “CoordinateCleaner” package^89^. For each species, we buffered each of its occurrence points by five kilometers and calculated the percent of buffered points that overlapped the species’ AOH.

### 3. Estimation of bat-human overlap, bat species richness, and habitat proportions

To identify regions where human populations might be exposed directly or indirectly to SARSr-CoV bat hosts, we overlaid human population count data on each host species’ AOH. We used a 1-km resolution raster of 2020 population count data from WorldPop^90^ and resampled it with bilinear interpolation using the “gdalUtils” package^91^ so that its extent and resolution matched that of the habitat raster. We calculated the number of people living within the AOH of each species separately, and divided this by the size of each AOH to obtain average human population density values. As living in areas with high diversity of SARSr-CoV bat hosts and large human populations may increase the likelihood of human-bat contact, viral spillover, and subsequent pathogen spread, we also visualized bat species richness and bat species richness multiplied by human population size (which we term “relative spillover risk”) across the consensus map. To examine the relative importance of different habitat types to SARSr-CoV bat hosts, we calculated the proportion (by area) of each habitat type within the AOH of each species.

### 4. Estimating human exposure to and infection with SARSr-CoVs

We used a probabilistic risk assessment to estimate the extent of SARSr-CoV spillover from bats to humans. We assumed that the number of people infected with SARSr-CoVs by bats each year is equal to 1) the number of people who live in the consensus area of SARSr-CoV bat hosts, multiplied by 2) the probability that a human comes into contact with a bat, multiplied by 3) the probability that a bat-human contact leads to a serologically detectable human infection, multiplied by 4) the probability that serological detection is due to an infection within the previous year. To inform our choice of distribution for each input variable, we gathered data from the literature on human-bat contacts, human SARSr-CoV seroprevalence, and human SARS antibody duration (see the Supplemental Material for details). We accounted for uncertainty and/or variation associated with the input variables by assigning a probability distribution (rather than a single fixed value) to each. We performed Latin hypercube sampling with the “lhs” package^92^ to generate 400,000 sets of input combinations and calculated the total number of people infected for each set of inputs, creating an output distribution. We then calculated summary statistics (mean, median, range) for this distribution. Finally, we performed a global sensitivity analysis by calculating Sobol sensitivity analyses to understand the relative contribution of each input variable to the outcome.

## Data Availability

Data Availability
The datasets analyzed during the current study are available on Zenodo: https://zenodo.org/record/5251726#.YSWivEApAUE.
Code Availability
Code to replicate the analyses and figures is available on Zenodo: https://zenodo.org/record/5251726#.YSWivEApAUE.

https://zenodo.org/record/5251726#.YSWivEApAUE

## Acknowledgments

We acknowledge funding from the National Institute of Allergy and Infectious Diseases of the National Institutes of Health (NIAID-CREID U01AI151797 “EID-SEARCH”), the Samuel Freeman Charitable Trust, Pamela Thye, The Wallace Fund, and an Anonymous Donor c/o Schwab Charitable. L.F.W. is supported by grants from NRF (NRF2012NRF-CRP001-056; NRF2016NRF-NSFC002-013), NMRC (MOH-OFIRG19MAY-0011; COVID19RF-003) and MOE (MOE2019-T2-2-130) of Singapore. The authors thank Cadhla Firth, Noam Ross, and Emma Mendelsohn for useful discussions.

## Data Availability

The datasets analyzed during the current study are available on Zenodo: https://zenodo.org/record/5251726#.YSWivEApAUE.

## Code Availability

Code to replicate the analyses and figures is available on Zenodo: https://zenodo.org/record/5251726#.YSWivEApAUE.

## Author Contributions

P.D. conceived of the study, C.A.S., H.L., K.L.P., C.Z.-T., and K.J.O. designed the methods, C.A.S. conducted the analyses, H.L. performed the literature search, C.A.S. and H.L. designed figures and tables, C.A.S. wrote the first draft of the manuscript, and all authors provided comments and edits to the manuscript throughout the writing process. All authors approved the submitted manuscript and agree to be held accountable for their own contributions and ensure that questions related to the accuracy or integrity of any part of the work are appropriately investigated, resolved, and the resolution documented in the literature.

