## Supplemental material for "A strategy to assess spillover risk of bat SARS-related coronaviruses in Southeast Asia"

**Methods**

*Habitat suitability*

Using the IUCN Red List data, we assessed if each of 22 habitat types was suitable for each bat host species. These included various forest habitats, shrublands, rocky areas and caves, and artificial habitats, chosen from a larger list of ~100 habitat types^1^ as being influential in determining the occurrence of one or more target host species (Table S1). A global land cover map of these habitat types^2,3^ included 18 of the 22 target habitats. Because two missing habitat types (*7.1. Caves and Subterranean Habitats (non-aquatic) – Caves* and *7.2. Caves and Subterranean Habitats (non-aquatic) – Other subterranean habitats*) were suitable for many host species, we supplemented the land cover dataset with the World Map of Carbonate Rock Outcrops v3.0^4^ as a proxy for karst landscape and cave distribution. This dataset comprises two layers, one with areas of continuous carbonate rocks and another with abundant but not continuous rocks; we used the former for our analyses. We rasterized the carbonate rock outcrop shapefile using the “fasterize” package^5^ and combined this with the land cover raster. When restricting species IUCN ranges by suitable habitat, we used the combined land cover/carbonate rock raster if a species was found in habitat type 7.1, and the land cover-only raster if a species was not found in this habitat type. The other two habitat types not represented on the land cover raster were only suitable for 1-2 host species (i.e. *14.6. Subtropical/Tropical Heavily Degraded Former Forest* and *15.8. Seasonally Flooded Agricultural Land*); therefore, we did not attempt to find other data to use as a proxy for these habitats.

*Extrapolating SARSr-CoV spillover in Southeast Asia*

We used a probabilistic risk assessment to estimate the extent of SARSr-CoV spillover from bats to humans. We assumed that the number of people infected with SARSr-CoVs by bats each year (*I_total_*) can be approximated as the number of people who live in the consensus area of SARSr-CoV bat hosts (*N_people_*), multiplied by the probability that a human comes into contact with a bat (*P_contact_*), multiplied by the probability that a bat-human contact leads to a serologically detectable human infection (*P_detection_*), multiplied by the probability that serological detection is due to an infection within the previous year (*P_pastyear_*) (Equation 1).

Eq. 1. *I_total_* = *N_people_* * *P_contact_* * *P_detection_* * *P_pastyear_*

We accounted for uncertainty and/or variation associated with the input variables by assigning a probability distribution (rather than a single fixed value) to each. We performed Latin hypercube sampling with the “lhs” package^6^ to generate 400,000 sets of input combinations and calculated *I_total_* for each set of inputs, creating a distribution for *I_total_*. We then calculated summary statistics (mean, median, range) for this distribution. Our choices of probability distributions for *N_people_*, *P_contact_*, *P_detection_*, and *P_pastyear_* are described below.

For *N_people_*, we assigned the normal distribution $N\left( 477589486, {15919650}^{2} \right)$. The mean value was derived from our calculation of the number of people living in the consensus area of SARSr-CoV bat hosts within Southeast Asia, based on 2020 WorldPop population count data. The standard deviation was chosen so that the tails of the distribution (3 standard deviations) would extend 10% beyond the mean, to reflect potential uncertainty associated with the WorldPop dataset.

To inform our choice of distribution for *P_contact_*, we gathered data on bat-human contacts in Southeast Asia using a systematic search of Google Scholar, PubMed, and Web of Science with the following keywords: ("bat contact" OR "human bat contact" OR "human bat interaction" OR "bat human contact" OR "bat human interaction") AND ("Bangladesh” OR “Bhutan” OR “Brunei” OR “Cambodia” OR “China” OR “India” OR “Indonesia” OR “Lao PDR” OR “Malaysia” OR “Myanmar” OR “Nepal” OR “Philippines” OR “Singapore” OR “Sri Lanka” OR “Thailand” OR “Timor-Leste” OR “Vietnam”). Only English language papers published from January 1, 2000 to May 07, 2021 were included in the search. A total of 421 records were initially identified, of which 24 were retained following title and abstract screening that excluded duplicate articles, studies that only focused on bat-borne pathogens or bat ecology, and studies based outside of Southeast Asia. Full text review was performed on these 24 articles. Six articles provided data on the frequency of human contact with bats or their excreta within the last 12 months. These studies included specific data on behaviors and practices that might allow viral transmission (e.g. eating bats, receiving a bat scratch or bite, exposure to urine or guano, being in a bat cave, having bats in one’s house; summarized in Table S3). Given that the probability of contact is bounded between 0 and 1, we assigned the beta distribution $\beta\left( 0.9366017, 5.2604551 \right)$ to these data, using the “fitdistrplus” package^7^ to determine the shape parameters.

In a similar manner, to inform our choice of distribution for *P_detect_*, we gathered data on viral seroprevalence among people with self-reported bat contact using a systematic search of Google Scholar, PubMed, and Web of Science with the following keywords: ("bat contact" OR "human bat contact" OR "human bat interaction" OR "bat human contact" OR "bat human interaction") AND ("serological prevalence” OR “seroprevalence” OR “serological evidence” OR “human infection” OR “spillover”). Only English language papers published from January 1, 2000 to June 21, 2021 were included in the search. A total of 339 records were initially identified, of which 12 were retained following title and abstract screening that excluded i) duplicate articles, ii) studies that reported seroprevalence in animal populations, iii) studies that provided either seroprevalence or human-bat contact but not both, iv) studies that detected pathogens that are unlikely to be transmitted from contact with bats, and v) studies that analyzed human-bat contact only among identified seropositive human cases. Full text review was performed on the remaining 12 articles. Seven articles provided data on human viral seroprevalence among populations who reported contact with bats (Table S4). We assigned the beta distribution $\beta\left( 0.2941532, 13.7297947 \right)$ to these data, again using the “fitdistrplus” package^7^ to determine the shape parameters. We note that it is unlikely that all known and as-yet-undiscovered bat SARSr-CoVs are likely able to infect people directly, or at all. However, the use of serology data from human surveys provides a way to account for this by providing information on exposure that has led to prior infection, albeit that the severity of that infection remains unknown.

Studies of SARS patients have provided a range of estimates for the persistence of antibodies at diagnostically detectable levels, with a maximum of 6 years, suggesting that the proportion of those testing seropositive could include people infected more than one year prior to testing. We note that studies of memory T cells have shown that it is possible to identify antibodies as long as 17 years after SARS infection^8^. However, our analysis uses published data on serological tests, not memory T-cell activation studies, therefore we set the maximum at 6 years, which is the longest time after exposure that published data for serological testing provided evidence of detectable IgG. Given that our aim was to estimate the amount of bat-to-human spillover each year, we performed a calculation to estimate the likelihood that someone who tested seropositive was infected within the past year. We gathered data from papers that reported the duration of SARS-CoV immunoglobulin G (IgG) antibody detection among patients who recovered from SARS (Table S5). Generally, the detectability of IgG rose rapidly following infection, peaked at 3-4 months following symptom onset and remained high for the first 16 months following symptom onset. Detectability then decreased over time, with ~40-70% of patients having detectable IgG after three years, and only ~10% having detectable IgG after six years. We fit a 2nd order polynomial to these data, then divided the integral from 0-12 months by the integral from 0-71.5 months (when antibody detection declines to ~0 on the curve) to calculate the probability that antibody detection is due to an infection in the past year. We then repeated this process excluding the only data point at 72 months, this time dividing the integral from 0-12 months by the integral from 0-49.5 months (when antibody detection goes to ~0 on the curve). We used the two values generated from these processes to assign the uniform distribution $\beta\left( 0.232444, 0.3059904 \right)$ for *P_pastyear_*.

Finally, we performed a global sensitivity analysis by calculating first-order and total-order Sobol sensitivity indices^9^ for the four input variables. Sobol sensitivity indices measure the amount of variance in an outcome (here, the total number of people in Southeast Asia infected each year by SARSr-CoVs by bat hosts) due to input variables, and can be used for non-linear relationships. The first-order index represents the amount of variance in the outcome due to each input on its own. The total-order index represents the amount of variance in the outcome due to each input including interactions with other inputs. First-order and total-order indices are always ≤ 1, and first-order indices are ≤ total-order indices. All sensitivity indices were calculated using the “sensitivity” package^10^ using the Janon-Monod method^11,12^.

**Results**

Our Latin hypercube sampling, run over 400,000 trials, produced a right-skewed distribution for the total number of people infected by SARSr-CoVs by bat hosts each year in Southeast Asia (Fig. S3). The mean and median values of this distribution were 407,422 and 53,225, respectively, while the range of the distribution was 1 - 35,632,228. Sensitivity analyses (Fig. S4) indicated that the probability of antibody detection given contact with a bat contributed most to variance in the outcome (first-order sensitivity index: 0.486; 95% CI: 0.475-0.496; total-order sensitivity index: 0.873; 95% CI: 0.864-0.881). The probability of contact with a bat contributed moderately to variance in the outcome (first-order sensitivity index: 0.125; 95% CI: 0.117-0.134; total-order sensitivity index: 0.509; 95% CI: 0.499-0.519). The number of people in the consensus area of SARSr-CoV bat hosts, as well as the probability that IgG detection was due to an infection in the past year, contributed little to outcome variance (Fig. S4).

**Table S1.** A compiled list of bat SARSr-CoV hosts, their elevation limits, and the suitability of selected IUCN habitat types. An **X** indicates that a habitat is suitable for a species. Gray shading of cells indicates habitat types not included in a global map of terrestrial habitat types^2,3^. All elevation and suitability data were sourced from IUCN^13^.

| **Species** | **Family** | **Reference** | **Elevation limits (m)** | | **Habitat type*** | | | | | | | | | | | | | | | | | | | |
| --- | --- | --- | --- | --- | --- | --- | --- | --- | --- | --- | --- | --- | --- | --- | --- | --- | --- | --- | --- | --- | --- | --- | --- | --- |
|  |  |  | **Upper** | **Lower** | **1.4** | **1.5** | **1.6** | **1.9** | **3.4** | **3.5** | **3.7** | **3.8** | **4.4** | **6.0** | **7.1** | **7.2** | **8.2** | **14.1** | **14.2** | **14.3** | **14.4** | **14.5** | **14.6** | **15.8** |
| *Aselliscus stoliczkanus* | Hipposideridae | ^14^ | NA | NA |  |  |  |  |  |  |  |  |  |  | **X** |  |  |  |  |  |  |  |  |  |
| *Chaerephon plicatus* | Molossidae | ^15^ | 950 | 0 |  | **X** | **X** |  |  |  |  |  |  |  | **X** | **X** |  |  | **X** |  | **X** |  |  | **X** |
| *Hipposideros armiger* | Hipposideridae | ^16^ | 2031 | 100 |  |  |  | **X** |  |  |  |  |  |  | **X** | **X** |  |  |  |  |  |  |  |  |
| *H. galeritus* | Hipposideridae | ^17^ | 1100 | 0 |  | **X** | **X** |  |  |  |  |  |  | **X** | **X** | **X** |  |  |  |  |  |  |  |  |
| *H. gentilis*** | Hipposideridae | ^16^ | 1900 | 0 |  |  | **X** | **X** |  |  |  |  |  |  | **X** | **X** |  |  |  |  |  |  |  |  |
| *H. larvatus* | Hipposideridae | ^17^ | 2000 | 0 |  |  | **X** | **X** |  |  |  |  |  |  | **X** | **X** |  |  |  |  |  |  |  |  |
| *H. pratti* | Hipposideridae | ^16^ | 2000 | 100 |  |  |  |  |  |  |  |  |  |  | **X** |  |  |  |  |  |  |  |  |  |
| *Nyctalus leisleri* | Vespertilionidae | ^18^ | 2400 | 0 | **X** |  |  |  |  |  |  | **X** |  | **X** |  |  |  |  | **X** |  |  |  |  |  |
| *Rhinolophus acuminatus* | Rhinolophidae | ^19^ | 1676 | 60 |  |  | **X** |  |  |  |  |  |  |  | **X** |  |  |  |  |  |  |  |  |  |
| *R. affinis* | Rhinolophidae | ^20^ | 2000 | 290 |  | **X** | **X** | **X** |  |  |  |  |  |  | **X** |  |  |  |  |  | **X** |  |  |  |
| *R. creaghi* | Rhinolophidae | ^17^ | 1500 | 0 |  |  | **X** |  |  |  |  |  |  |  | **X** |  |  |  |  |  |  |  |  |  |
| *R. ferrumequinum* | Rhinolophidae | ^17^ | 3000 | 0 | **X** |  |  |  |  |  |  | **X** | **X** |  | **X** | **X** |  |  | **X** |  |  |  |  |  |
| *R. hipposideros* | Rhinolophidae | ^21^ | 2000 | 0 | **X** |  |  |  | **X** | **X** | **X** | **X** | **X** |  | **X** | **X** |  |  |  |  |  | **X** |  |  |
| *R. macrotis* | Rhinolophidae | ^14^ | 1692 | 200 |  | **X** | **X** | **X** |  |  |  |  |  |  | **X** | **X** |  |  |  |  |  |  |  |  |
| *R. malayanus* | Rhinolophidae | ^22^ | 1400 | 0 |  |  | **X** |  |  |  |  |  |  |  | **X** |  |  | **X** |  | **X** | **X** |  |  |  |
| *R. pearsonii* | Rhinolophidae | ^14^ | 3077 | 123 | **X** |  | **X** | **X** |  |  |  |  |  |  | **X** | **X** |  |  |  |  |  |  |  |  |
| *R. pusillus* | Rhinolophidae | ^14^ | 1370 | 200 |  | **X** | **X** |  |  |  |  |  |  |  | **X** | **X** |  |  |  |  |  |  |  |  |
| *R. rex* | Rhinolophidae | ^23^ | NA | NA |  |  | **X** |  |  |  |  |  |  |  | **X** |  |  |  |  |  |  |  |  |  |
| *R. shameli* | Rhinolophidae | ^24^ | NA | NA |  | **X** | **X** |  |  |  |  |  |  |  | **X** |  |  |  |  |  |  |  |  |  |
| *R. sinicus* | Rhinolophidae | ^17^ | 2769 | 500 | **X** | **X** | **X** |  |  |  |  |  |  |  | **X** | **X** |  |  |  |  |  |  |  |  |
| *R. stheno* | Rhinolophidae | ^25^ | 1700 | 0 |  | **X** |  |  |  |  |  |  |  |  |  |  |  | **X** |  | **X** |  |  | **X** |  |
| *R. thomasi* | Rhinolophidae | ^16^ | 1100 | 400 |  |  |  |  |  |  |  |  |  |  | **X** |  |  |  |  | **X** | **X** |  |  |  |
| *Tadarida teniotis* | Molossidae | ^26^ | 3100 | 0 |  |  |  |  | **X** |  |  | **X** | **X** | **X** | **X** | **X** | **X** |  |  |  |  | **X** |  |  |

*Habitat type names: 1.4. Forest -- Temperate; 1.5. Forest -- Subtropical/tropical dry; 1.6. Forest -- Subtropical/tropical moist lowland; 1.9. Forest -- Subtropical/tropical moist montane; 3.4. Shrubland -- Temperate; 3.5. Shrubland -- Subtropical/tropical dry; 3.7. Shrubland -- Subtropical/tropical high altitude; 3.8. Shrubland -- Mediterranean-type shrubby vegetation; 4.4. Grassland -- Temperate; 6.0. Rocky areas (e.g. inland cliffs, mountain peaks); 7.1. Caves and Subterranean Habitats (non-aquatic) -- Caves; 7.2. Caves and Subterranean Habitats (non-aquatic) -- Other subterranean habitat; 8.2. Desert -- Temperate; 14.1. Arable Land; 14.2. Pastureland; 14.3. Plantations; 14.4. Rural Gardens; 14.5. Urban Areas; 14.6. Subtropical/Tropical Heavily Degraded Former Forest; 15.8. Seasonally Flooded Agricultural Land

***H. gentilis* was listed as *H. pomona* in Latinne et al. 2020, but *H. gentilis* was used for our analyses due to recent taxonomic revisions; see Methods.

**Table S2.** Validation of species AOHs using cleaned occurrence records from the Global Biodiversity Information Facility (GBIF). Species are listed in order of decreasing percent of points within 5 km of AOH.

| **Species** | **Number of cleaned GBIF occurrence points** | **Number (%) of points within 5 km of AOH** |
| --- | --- | --- |
| *Rhinolophus creaghi* | 31 | 31 (100) |
| *R. malayanus* | 8 | 8 (100) |
| *Chaerephon plicatus* | 53 | 45 (85) |
| *Hipposideros galeritus* | 44 | 37 (84) |
| *H. larvatus* | 49 | 41 (84) |
| *R. shameli* | 9 | 7 (78) |
| *R. pearsonii* | 22 | 17 (77) |
| *R. macrotis* | 3 | 2 (67) |
| *R. thomasi* | 6 | 4 (67) |
| *R. affinis* | 74 | 48 (65) |
| *H. gentilis* | 29 | 18 (62) |
| *R. pusillus* | 36 | 22 (61) |
| *R. stheno* | 10 | 6 (60) |
| *Aselliscus stoliczkanus* | 6 | 3 (50) |
| *H. armiger* | 179 | 58 (32) |
| *R. acuminatus* | 14 | 3 (21) |
| *R. sinicus* | 12 | 2 (17) |
| *R. rex* | 6 | 0 (0) |
| *Tadarida teniotis* | 2 | 0 (0) |
| *Nyctalus leisleri* | 1 | 0 (0) |
| *H. pratti* | 0 | NA |
| *R. ferrumequinum* | 0 | NA |
| *R. hipposideros* | 0 | NA |

**Table S3.** Prevalence of human-bat contact within countries in Southeast Asia. Studies were identified via a systematic literature search (see Supplemental Methods).

| **Country** | **Study population** | **Description of bat contact** | **Prevalence of bat contact (n/X)** | **Timeframe of contact with bat** | **Reference** |
| --- | --- | --- | --- | --- | --- |
| China | People living in areas near bat populations | Bats in house | 12.7% (201/1585) | Previous 12 months | ^27^ |
|  |  | Cooked or handled bats | 0.6% (9/1585) |  |  |
| Indonesia | People (mostly men) living in/near a forest conservation area | Expelling | 10.7% (16/150) | Not reported | ^28^ |
|  |  | Hold/capture/hunt | 46.0% (69/150) |  |  |
|  |  | Cutting | 13.3% (20/150) |  |  |
|  |  | Cooking | 13.3% (20/150) |  |  |
|  |  | Eating | 16.7% (25/150) |  |  |
|  |  | Selling | 3.3% (5/150) |  |  |
| Thailand | People living in areas with high bat density | Found live bat(s) in house, community, or tourist location | 20.4% (128/626) | Previous 6 months | ^29^ |
|  |  | Consumed bat meat | 15.3% (96/626) |  |  |
|  |  | Cleaned bat guano from house or the community | 12.5% (78/626) |  |  |
|  |  | Found dead bat(s) in house | 10.4% (65/626) |  |  |
|  |  | Bat guano mining/collecting | 7.3% (46/626) |  |  |
|  |  | Cleaned bat carcasses from house or the community | 7.2% (45/626) |  |  |
|  |  | Other activities (e.g. hunted bats, exposed to bat urine) | 6.5% (41/626) |  |  |
|  |  | Used bat guano | 4.0% (25/626) |  |  |
|  |  | Bitten by a bat | 2.6% (16/626) |  |  |
|  | Adult guano miners, bat hunters, game wardens, residents/personnel at temples with large bat roosts | Inside bat cave or roost area | 57.5% (61/106) | Exposed > 5 times/year | ^30^ |
|  |  | Direct bat contact (unspecified) | 27.4% (29/106) |  |  |
|  |  | Bat consumption | 10.4% (11/106) |  |  |
|  |  | Bat scratch | 5.7% (6/106) |  |  |
|  |  | Bat bite | 1.9% (2/106) |  |  |
|  | People living in areas where bat roosts are present | Various (hunting bats, eating bats, collecting bat guano, cleaning bat feces, finding bat carcasses in houses and communities) | 46.6% (142/305) | Previous 6 months | ^31^ |
| Vietnam | People living in farming communities involved in raising, slaughtering, or processing wildlife or livestock | Maintained bat roosts to produce guano for fertilizer | 1.2% (3/245) | Previous 12 months | ^32^ |

**Table S4.** Human viral seroprevalence among individuals reporting contact with bats. Studies were identified via a systematic literature search (see Supplemental Methods).

| **Country** | **Pathogen** | **Description of bat contact** | **Seroprevalence (X/n)** | **Reference** |
| --- | --- | --- | --- | --- |
| Cameroon | Nipah virus | Hunting bats | 3.0% (3/99) | ^33^ |
|  |  | Butchering bats | 4.1% (7/171) |  |
|  |  | General contact with bats (including hunting, butchering, and receiving bite or scratch) | 3.1% (7/227) |  |
| Cambodia | Nipah virus | Hunting bats | 0% (0/4) | ^34^ |
|  |  | Palm-juice collection and selling | 0% (0/15) |  |
| Australia | Hendra virus (then named equine morbillivirus) | “Prolonged, significant contact” in the context of being a flying fox carer | 0% (0/128) | ^35^ |
| China | SARSr-CoV | General contact with bats | 0.5% (1/199) | ^27^ |
|  | HKU10-CoV | General contact with bats | 0.5% (1/199) |  |
|  | HKU9-CoV | General contact with bats | 0% (0/199) |  |
|  | MERS-CoV | General contact with bats | 0% (0/199) |  |
| Republic of Congo | Ebola virus | General exposure to bats | 14% (7/50) | ^36^ |
|  |  | Consumption of bats | 0% (0/23) |  |
| Malaysia | Tioman virus | Consumed fruit partially eaten by bats | 6.3% (2/32) | ^37^ |
| Malaysia | Nipah virus | Physical contact (unspecified) | 0% (0/15) | ^38^ |
|  |  | Consumed fruit partially eaten by bats | 0% (0/29) |  |

**Table S5.** Duration of SARS-CoV immunoglobulin G (IgG) antibody detection among patients who recovered from SARS

| **Time post-disease onset (as originally reported)** | **Time post-disease onset in months (used for curve fitting)** | **% (n/X) with detectable IgG** | **Reference** |
| --- | --- | --- | --- |
| 0-7 days | 0.25 | 11.8 (2/17) | ^39^ |
| 8-14 days | 0.5 | 38.5 (10/26) | ^39^ |
| 15-20 days | 0.75 | 77.3 (17/22) | ^39^ |
| 21-30 days | 1 | 91.7 (33/36) | ^39^ |
| 1 month | 1 | 100 (36/36) | ^40^ |
| 1 month | 1 | 100 (17/17) | ^41^ |
| 1 month | 1 | 100 (37/37) | ^42^ |
| 31-60 days | 2 | 93.1 (67/72) | ^39^ |
| 61-90 days | 3 | 94.3 (33/35) | ^39^ |
| 3 months | 3 | 100 (19/19) | ^43^ |
| 4 months | 4 | 100 (36/36) | ^40^ |
| 4 months | 4 | 100 (41/41) | ^42^ |
| 91-120 days | 4 | 100 (11/11) | ^39^ |
| 5 months | 5 | 100 (13/13) | ^44^ |
| 7 months | 7 | 100 (41/41) | ^40^ |
| 7 months | 7 | 100 (44/44) | ^42^ |
| 121-210 days | 7 | 100 (23/23) | ^39^ |
| 10 months | 10 | 100 (37/37) | ^40^ |
| 10 months | 10 | 100 (37/37) | ^42^ |
| 12 months | 12 | 94.7 (18/19) | ^43^ |
| 1 year | 12 | 100 (17/17) | ^41^ |
| 211-365 days | 12 | 93.9 (46/49) | ^39^ |
| 16 months | 16 | 100 (32/32) | ^40^ |
| 16 months | 16 | 100 (32/32) | ^42^ |
| 18 months | 18 | 84.2 (16/19) | ^43^ |
| 20 months | 20 | 82.4 (14/17) | ^44^ |
| 24 months | 24 | 88.2 (30/34) | ^40^ |
| 24 months | 24 | 84.2 (16/19) | ^43^ |
| 24 months | 24 | 88.6 (31/35) | ^42^ |
| 366-763 days | 24 | 89.6 (86/96) | ^39^ |
| 30 months | 30 | 80.6 (29/36) | ^42^ |
| 35 months | 35 | 84.6 (11/13) | ^44^ |
| 36 months | 36 | 74.2 (23/31) | ^42^ |
| 36 months | 36 | 42.1 (8/19) | ^43^ |
| 764-1265 days | 36 | 53.6 (15/28) | ^39^ |
| 72 months | 72 | 8.9 (2/23) | ^45^ |

**Figure S1.** Comparison of original IUCN species range sizes to AOH sizes.

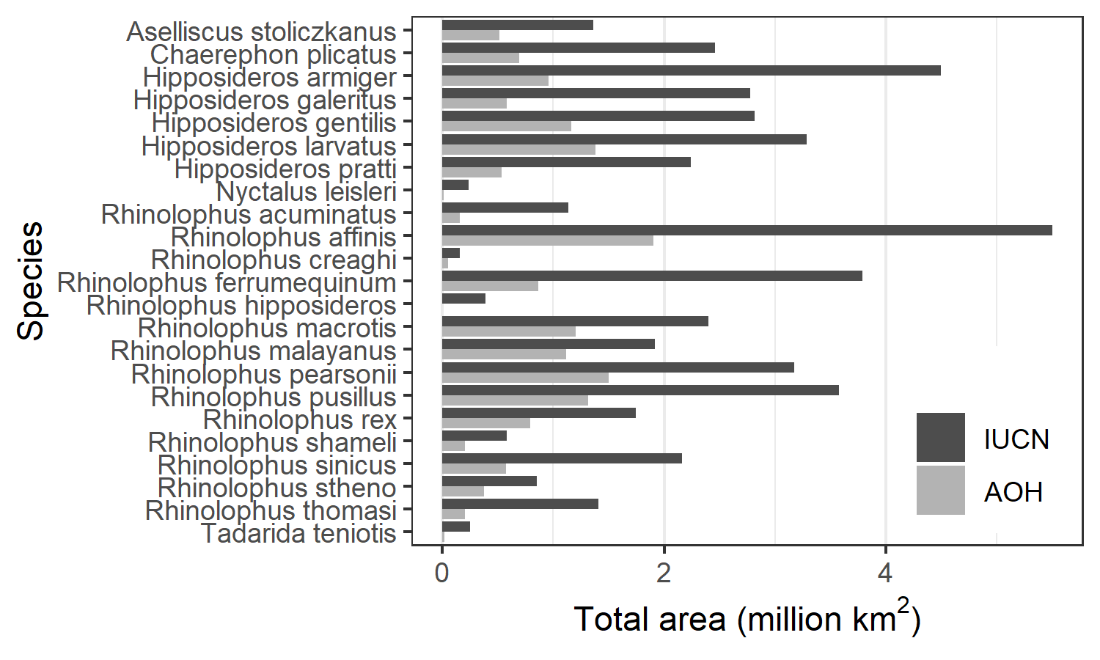

**Figure S2.** Human population density per km^2^ within the AOH of each SARSr-CoV bat host species

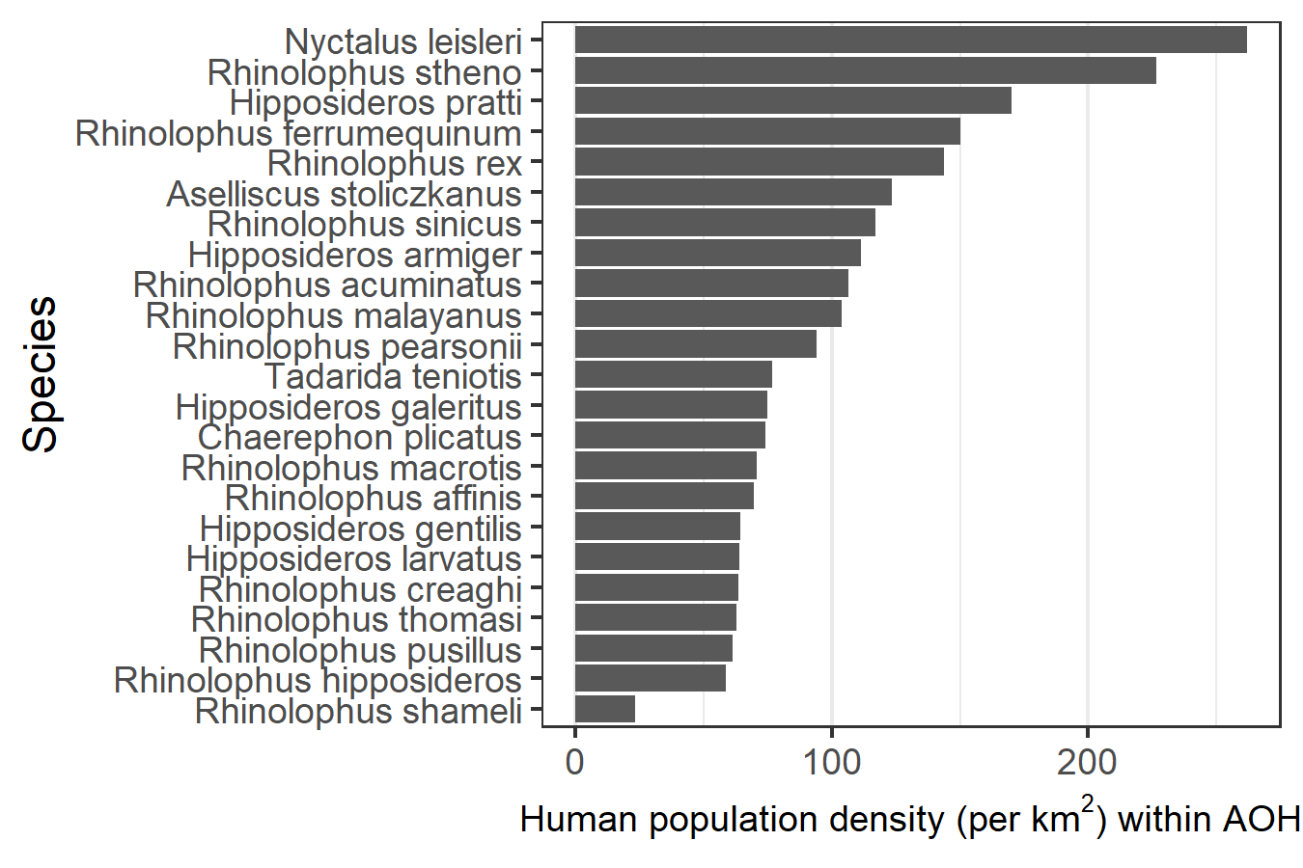

**Figure S3.** Histograms of input variables generated with Latin hypercube sampling (from left to right: number of people in bat SARSr-CoV host range, probability of contact with bat, probability of IgG detection given contact with bat, probability that IgG detection is due to an infection in the past year) and output (total people infected).

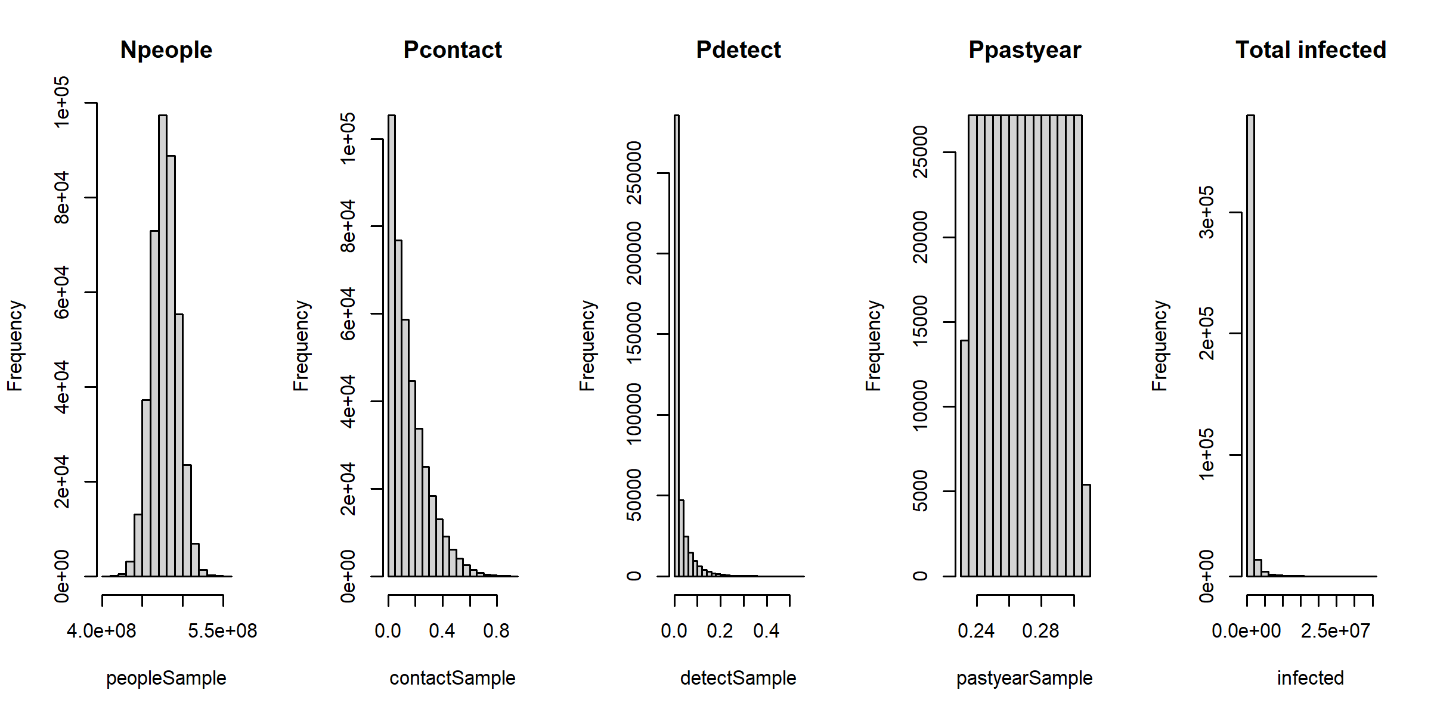

**Figure S4.** **Top row:** Estimated total number of people in Southeast Asia infected with SARSr-CoVs by bats each year, plotted as a function of four input variables. **Bottom row:** Sobol sensitivity indices, indicating the amount of variance in the outcome due to each input on its own (first-order) and the amount of variance in the outcome due to each input including interactions with other inputs (total-order). Error bars represent 95% confidence intervals.

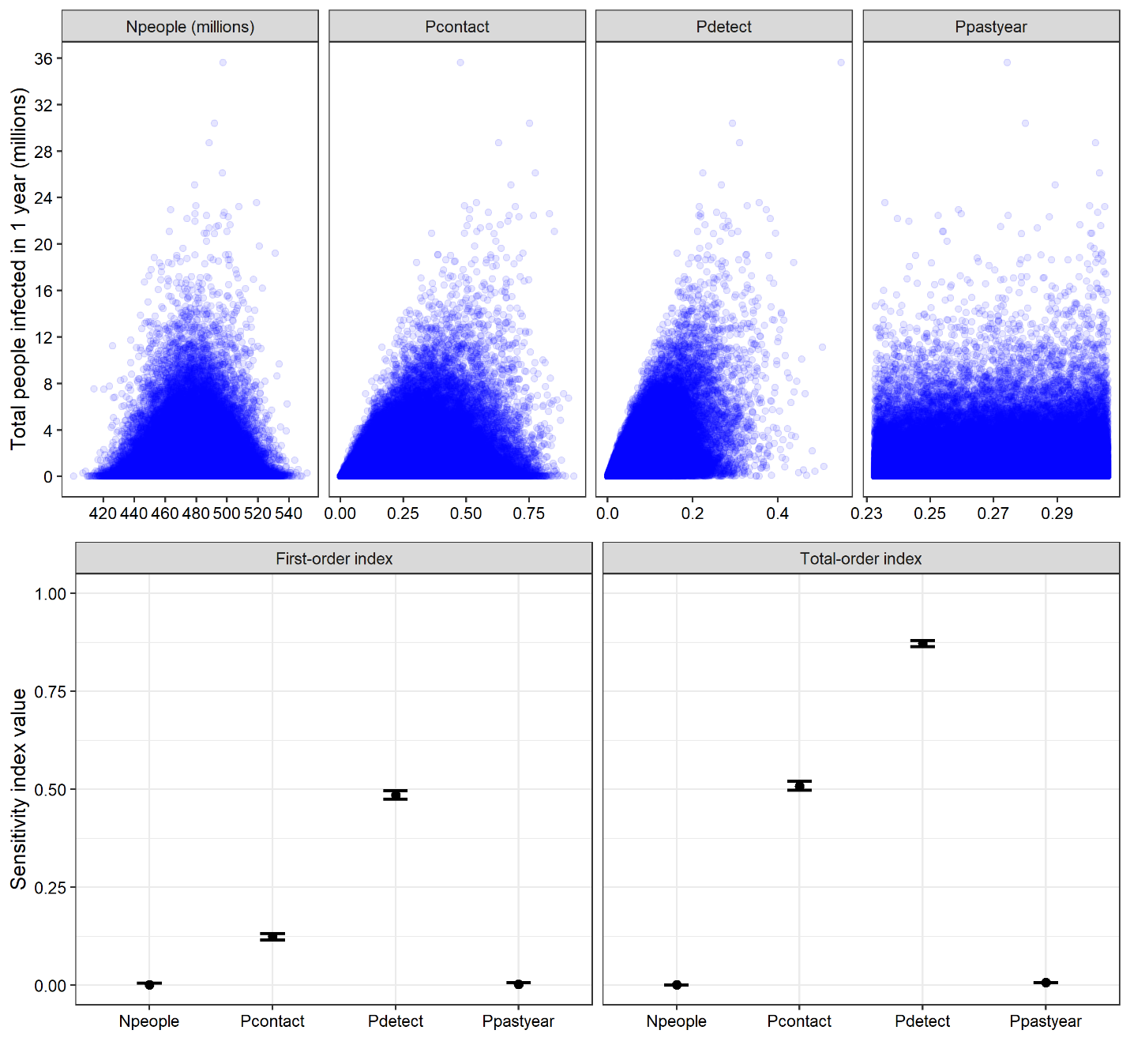
